# C-Reactive protein and SOFA score as early predictors of critical care requirement in patients with COVID-19 pneumonia in Spain

**DOI:** 10.1101/2020.05.22.20110429

**Authors:** Luis Mario Vaquero-Roncero, Elisa Sánchez-Barrado, Daniel Escobar-Macias, Pilar Arribas-Pérez, Jose Ramón Gonzalez-Porras, Jesús F Bermejo-Martín, Cristina Doncel, JM Bastida, Azucena Hernández-Valero, Carolina Jambrina-García Montoto, José Sánchez-Crespo, Pablo Alonso-Hernández, Domingo Bustos-García, Antonio Rodríguez-Calvo, Gerardo Riesco-Galache, Raúl Alzibeíro, Alberto Hernández-Sánchez, Rocío Eiros, M Carmen Vargas, María Ángeles Martín, Teresa López, José Alfonso Sastre, José Carlos Garzón, Mar Fernández, Belén García, M Magdalena Muñoz, María Isabel Martínez, Gonzalo García, Agustín Díaz, Edgar Marcano, Víctor Sagredo-Meneses, Miguel Vicente Sánchez-Hernandez

## Abstract

**Background:** Some patients infected by SARS-CoV-2 in the recent pandemic have required critical care, becoming one of the main limitations of the health systems. Our objective has been to identify potential markers at admission predicting the need for critical care in patients with COVID-19 pneumonia.

**Methods:** We retrospectively collected and analyzed data from electronic medical records of patients with laboratory-confirmed SARS-CoV-19 infection by real-time RT-PCR. A comparison was made between patients staying in the hospitalization ward with those who required critical care. Univariable and multivariable logistic regression methods were used to identify risk factors predicting critical care need.

**Findings:** Between March 15 and April 15, 2020, 150 patients under the age of 75 were selected (all with laboratory confirmed SARS-CoV-19 infection), 75 patients requiring intensive care assistance and 75 remaining the regular hospitalization ward. Most patients requiring critical care were males, 76% compared with 60% in the non-critical care group (p<0.05). Multivariable regression showed increasing odds of in-hospital critical care associated with increased C-reactive protein (CRP) (odds ratio 1.052 (1.009-1.101); p=0.0043) and higher Sequential Organ Failure Assessment (SOFA) score (1.968 (1.389-2.590) p<0.0001) both at the time of hospital admission. The AUC-ROC for the combined model was 0.83 (0.76-0.90) (vs AUC-ROC SOFA p<0.05).

**Interpretation:** Patients COVID-19 positive presenting at admission with high SOFA score ≥2 combined with CRP ≥ 9,1 mg/mL could help clinicians to identify them as a group that will more likely require critical care so further actions might be implemented to improve their prognosis.

## Introduction

In December 2019, an atypical pneumonia appears, which is later known to be caused by a novel coronarivus, (SARS-CoV-2). In April 2020 the virus has spread to 212 countries around the world, infecting more than 1,607,467 people and causing more than 98,866 deaths (1). On May 9 in Spain, 223,578 cases of infection were diagnosed with 26,478 related deaths (2). At the time of illness onset, patients may experience low-degree fever or flu-like symptoms, but suddenly, severe respiratory failure (SRF) may appear (3). Patients with SARS-CoV-2 infection have high rates of hospitalization and intensive care unit (ICU) admission (4). There have been a large number of parameters that have been related to morbidity and mortality (3,5,6,7,8), ranging from sex (9) to increased circulating levels of D-dimers (3) which may suggest endothelial activation. Furthermore, in most reports an association between severe clinical deterioration and a possible cytokine storm has been described, characterized by the release of cytokines like IL-6 and IL-1 but also classic markers of inflammation like C-reactive protein (CRP) and ferritin levels (10,11). Moreover, some reports suggest the possibility of different inflammation profiles predicting severity that could be identified in serum.

On the other hand, some patients infected by SARS-CoV-2 may need critical care, becoming one of the main limitations of the health systems in pandemic cases (12). To date, we do not know clearly which comorbidities, laboratory test results or severity characteristics of the patients are able to predict the potential need of these resources, resulting into the collapse of the healthcare system. Appropriate triage tools as well as the identification of patients’ profiles at high risk of severity would be crucial to individualize the management (13).

Based on this background, we decided to evaluate our series of patients COVID-19 to identify markers at the moment of admission at the hospital able to predict the need of critical care assistance.

## Material and methods

### Patient selection

We carried out a retrospective study collecting data from 150 patients diagnosed with COVID-19 pneumonia. All had a confirmed diagnosis of SARS-CoV-2. Seventy-five patients were randomly selected from those admitted to the critical care units within the University Hospital of Salamanca (Critical care group (CG)) between March 15 to April 15, 2020. From a previous pilot study with 10 patients, we knew that the age of those admitted to the critical care unit were less than 75 years and their Charlson comorbidity index (14) was less than 6, so in the same way, and in the same period of time, we selected 75 hospitalized patients who did not require critical care, aged with less than 75 years and with a Charlson comorbidity index equal to or less than 6 (Non-Critical care group (nCG)). This group represents our control group to be compared with the group that required admission into the critical care unit.

Patients aged less than 18 or older than 76 years and also people with Charlson comorbidity index of more than 6 or with laboratory test critical for the evaluation of severity missing were excluded.

The criteria for admission to the critical care unit included refractory severe respiratory failure secondary to COVID-19 pneumonia with or without respiratory distress according to reference studies(15).

This study was approved by the Research Ethics Commission of the clinical hospital of Salamanca (PI 2020 05 487) and the requirement for informed consent was waived by the Ethics Commission.

### Data collection

In each group, we collected all data from the medical records including clinical and anthropometric variables, as well as laboratory results upon arrival at the hospital emergency department, prior to admission. We recorded the symptomatology reported by patients and the onset of symptoms and in addition the date of admission at the hospital and at the critical care unit, as well as treatments received. The Sequential Organ Failure Assessment SOFA (SOFA or SOFA score) was collected as a severity scale upon arrival at the hospital (16). All data were checked by two physicians (PA and ESB) and a third researcher (LMVR) resolved the discordances.

CRP and ferritin were selected as biomarkers of the activation of different inflammation tracks which could be related with the clinical deterioration (10,11).Depending on their values in the blood test we created four inflammation profiles.

### Laboratory procedures

SARS-CoV-2 one-step real-time reverse transcriptase–polymerase chain reaction (rRT-PCR) diagnostic assay in a nasopharyngeal swab was performed in all patients at the moment of admission. Blood test analysis included complete blood cell count, coagulation profile, serum biochemical tests (including renal and liver function, creatine kinase, lactate dehydrogenase, and electrolytes), C-reactive protein (CRP) interleukin-6 (IL-6), serum ferritin, and procalcitonin. All patients had chest radiographs and Computerized Tomography (CT) scans if required during their stay.

### Definitions

Fever was defined as axillary temperature of at least 38°C. COVID-19 pneumonia was described as respiratory symptoms (fever, dry cough, dyspnea…) plus infiltrates on chest imaging (2,3).Acute respiratory distress syndrome (ARDS) was defined according to the interim guidance of WHO for novel coronavirus (15). Hypoxemia was defined as arterial oxygen tension (PaO_2_) over inspiratory oxygen fraction (FIO_2_) of less than 300 mm Hg (17) o SaO2 over FIO_2_ of less than 220 (18, 19). Severe hypoxemia was defined as PaO_2_/FIO_2_ less than 150 mm Hg (20).

### Statistical Analysis

A descriptive analysis was performed to summarize the data. No imputation was made for missing data. Normality of the continuous variables was evaluated with Kolmogorov–Smirnov test. The quantitative variables are expressed as mean and standard deviation or in median and interquartile range (IQ 25-75) and the qualitative values as percentages and absolute numbers. To compare quantitative variables, we used nonparametric tests if the distribution was not normal (Mann–Whitney U test) and parametric if it was normal (Student’s t test). Qualitative variables were compared using Chi square or Fisher’s exact test.

We calculated the areas under the curve (AUC-ROC) of the biomarkers with a p value <0.05 between the two groups in the univariate analysis. From those, we exclusively selected the four parameters with the best areas under the curve with the intention of including the least possible number of covariates for the sample size (21). The selected parameters will be the covariates in a multivariate model to predict the primary outcome. We excluded variables from the multivariable analysis if their between-group differences were not significant, if the number of events was too small to calculate odds ratios, and if they had collinearity with another variable.

Furthermore, for the selected variables, we calculated the cut-off points to differentiate between the two groups according to the best Youden index, calculating the sensitivity and specificity for those points. Next, a forward stepwise technique selection approach (logistic regression) is used to create a model for the selected biomarkers, requiring a p value of less than 0.05 for entering the model and p value of less than 0.10 to stay in the model. The model was performed using bootstrapped replicate dataset and average receiver operating curve AUC difference between these models and the best of biomarkers was calculated with 95% CIs. For this purpose, DeLong test was used.

Finally, we obtained cut-off points (CPm) of ferritin and CRP from the ROC curves of these variables according to the best Youden index, calculating the sensitivity and specificity for those points in order to define 4 inflammation profiles depending on the result of the laboratory test: profile 1 (if value CRP> CPm and ferritin> CPm), profile 2 (if value CRP> CPm and ferritin <CPm), profile 3 (if value CRP <cut CPm and ferritin> CPm) and profile 4 (CRP <CPm and ferritin <CPm), and we studied them in the two groups. Significance level was set at p <0.05. SPSS 21^®^ and Stata 15^®^ were used to perform the statistical analysis.

## Results

### Baseline characteristics of the patients

Between March 15^th^ and April 15^th^, 150 patients were included in this study: 75 patients who required admission at the Critical Care Group (CG) and 75 patients who did not and representing our control group. Four patients were excluded from the CG group because of some data were missing so the final sample size was 146 patients.

The baseline characteristics of the patients in both groups upon arrival at the hospital are represented in the Table 1. The median age of the patients was 66.00 (IQR 57.75-71) years. Overall, 68% patients were males (60% in non-critical care group (nCG) vs 76% in CG (p<0.05)).The most common underlying comorbidities were hypertension (62 [42.5%]) followed by obesity (46 [31.5%]), diabetes (31 [21.1%]) and cardiac disease (27 [18.5%]) and were balanced in both groups.

**Table 1.**
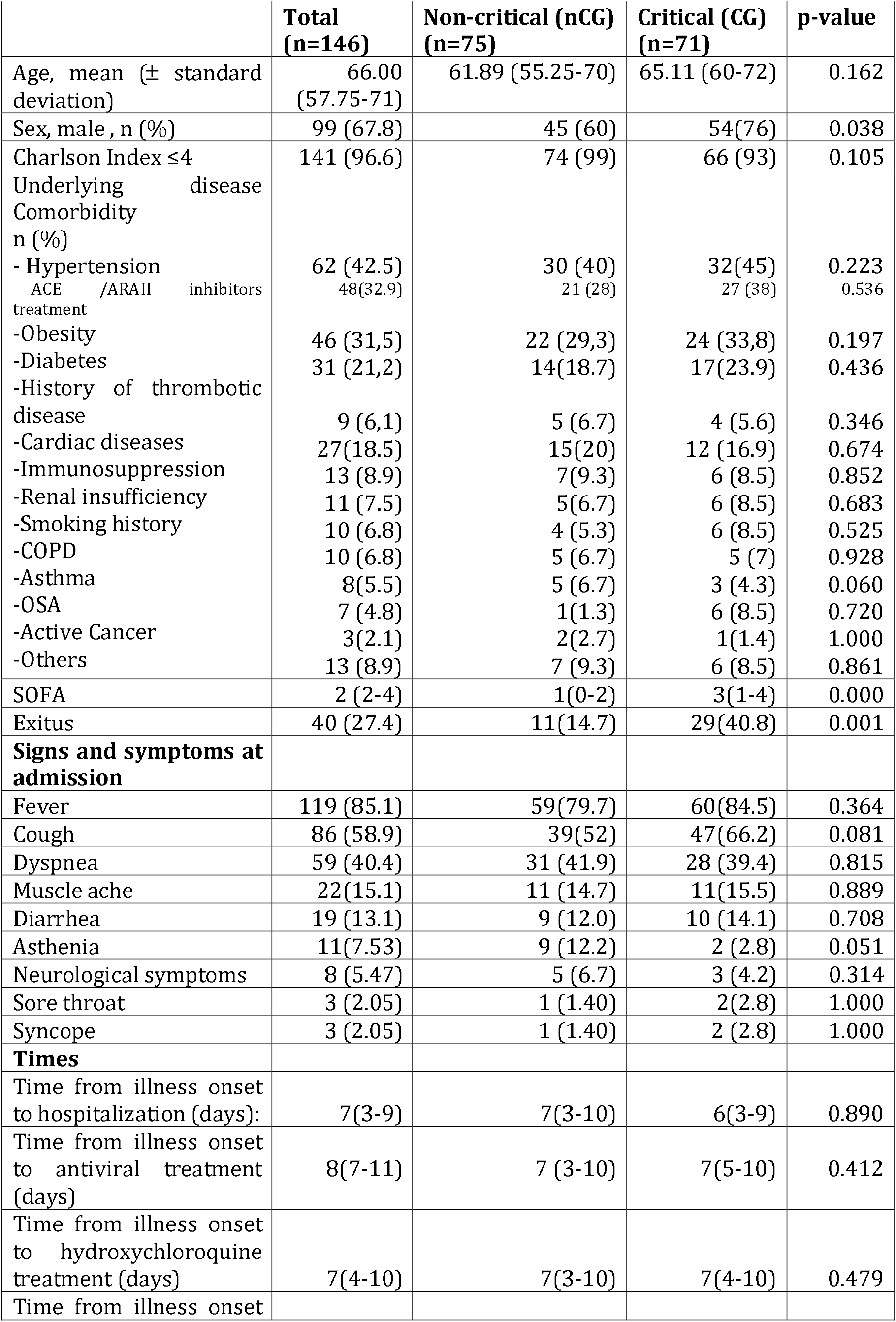
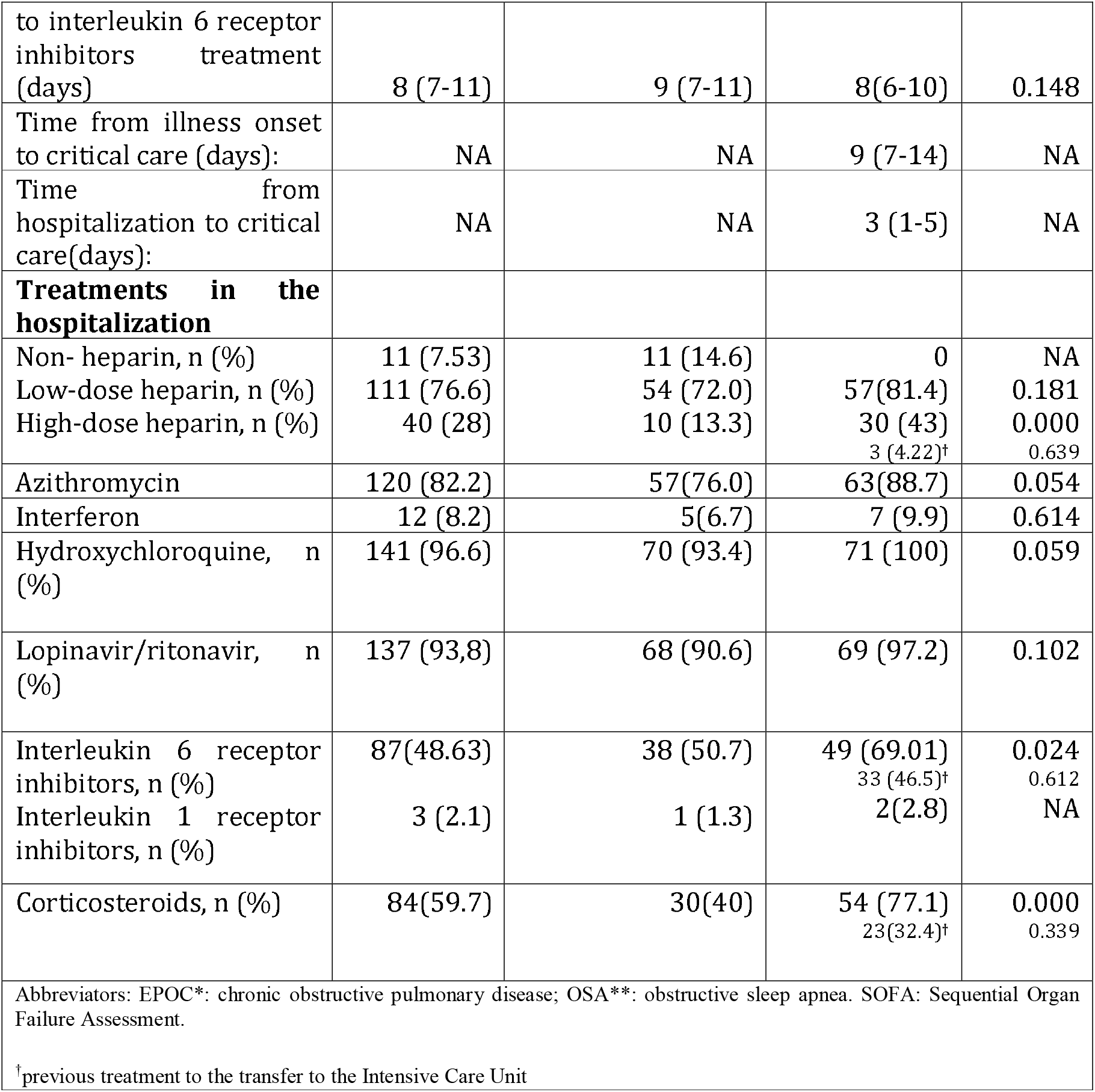
Baseline characteristic and parameters of COVID-19 patients on admission

Concerning symptoms and signs at the moment of admission at the hospital, fever (119 [85%] patients), cough (86 [59%]), and dyspnea (59 [40.4%]) were the most commonly reported without significant differences between both groups.

The median duration from onset of symptoms to admission at the hospital was 7.0 days (3.0–9.0), similar in non-critical and critical group. Time from illness onset to antiviral treatment was 8 days (7-11), similar in critical and non-critical. In the CG group, the median time from illness onset to admission in the critical care unit was 9 (7-14) days.

The inpatient mortality rate was 27.4% in the overall group and it was significantly higher in the CG group (40.8%) compared to the non-Critical group (14.7%).

The different treatments provided to patients during the hospitalization are represented in the table 1. Although there was observed a trend toward more patients in the CG receiving both hidroxicloroquine and azytromicine (p=0.05) the most significant differences were observed in the treatment with high-dose heparin, interleukine-6 receptor inhibitors and corticosteroids that were provided to 43%, 69% and 77.1%, of patients in the Critical group compared to 13.3% (p=0.000), 50.7% (p=0.024) and 40% (p=0.000) of patients in the non-critical group, respectively.

The table 2 represents the results of hemogram, coagulation and biochemistry analysis conducted at the moment of admission in the hospital, including pro-inflammatory markers. In the hemogram, higher level of neutrophils and lymphopenia were most frequently observed in the group of patients requiring admission into the critical care Units. Concerning biochemistry, higher levels of baseline creatinine, creatine kinase and lactate dehydrogenase were found in the CG group. In addition, patients requiring admission into the CG presented with higher levels of baseline fibrinogen as well as interleukine-6, C-reactive protein, procalcitonin and ferritin as pro-inflammatory markers. Finally, the SaO2/FiO2 values were significantly lower in critical than in non-critical patients group. Of note, patients that required admission at Critical Care Unit presented at admission with a significantly higher SOFA score (table 2).

**Table 2.**
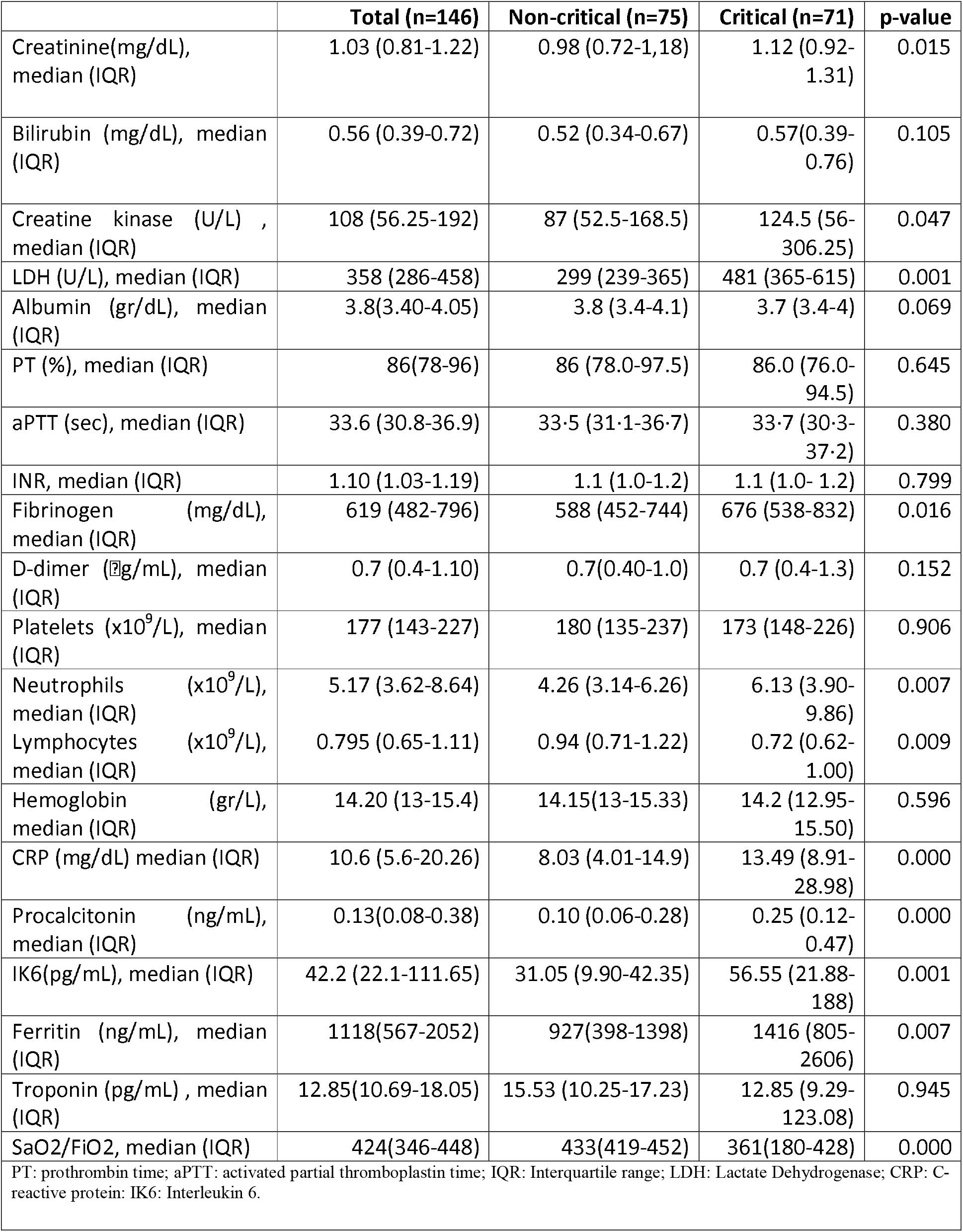
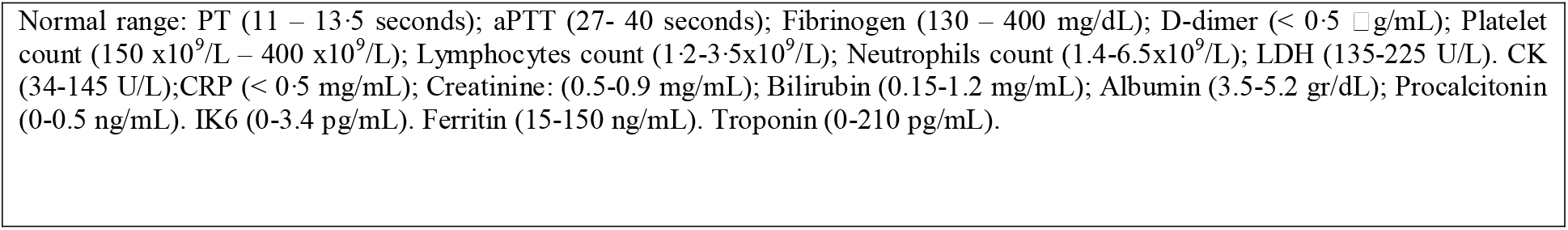
Analytical parameters of COVID-19 patients on hospital admission according to the place of subsequent admission.

|  | **Total (n=146)** | **Non-critical (n=75)** | **Critical (n=71)** | **p-value** |
| --- | --- | --- | --- | --- |
| Creatinine(mg/dL), median (IQR) | 1.03 (0.81-1.22) | 0.98 (0.72-1,18) | 1.12 (0.92-1.31) | 0.015 |
| Bilirubin (mg/dL), median (IQR) | 0.56 (0.39-0.72) | 0.52 (0.34-0.67) | 0.57(0.39-0.76) | 0.105 |
| Creatine kinase (U/L) , median (IQR) | 108 (56.25-192) | 87 (52.5-168.5) | 124.5 (56-306.25) | 0.047 |
| LDH (U/L), median (IQR) | 358 (286-458) | 299 (239-365) | 481 (365-615) | 0.001 |
| Albumin (gr/dL), median (IQR) | 3.8(3.40-4.05) | 3.8 (3.4-4.1) | 3.7 (3.4-4) | 0.069 |
| PT (%), median (IQR) | 86(78-96) | 86 (78.0-97.5) | 86.0 (76.0-94.5) | 0.645 |
| aPTT (sec), median (IQR) | 33.6 (30.8-36.9) | 33.5 (31.1-36.7) | 33.7 (30.3-37.2) | 0.380 |
| INR, median (IQR) | 1.10 (1.03-1.19) | 1.1 (1.0-1.2) | 1.1 (1.0- 1.2) | 0.799 |
| Fibrinogen (mg/dL), median (IQR) | 619 (482-796) | 588 (452-744) | 676 (538-832) | 0.016 |
| D-dimer (µg/mL), median (IQR) | 0.7 (0.4-1.10) | 0.7(0.40-1.0) | 0.7 (0.4-1.3) | 0.152 |
| Platelets (x10<sup>9</sup>/L), median (IQR) | 177 (143-227) | 180 (135-237) | 173 (148-226) | 0.906 |
| Neutrophils (x10<sup>9</sup>/L), median (IQR) | 5.17 (3.62-8.64) | 4.26 (3.14-6.26) | 6.13 (3.90-9.86) | 0.007 |
| Lymphocytes (x10<sup>9</sup>/L), median (IQR) | 0.795 (0.65-1.11) | 0.94 (0.71-1.22) | 0.72 (0.62-1.00) | 0.009 |
| Hemoglobin (gr/L), median (IQR) | 14.20 (13-15.4) | 14.15(13-15.33) | 14.2 (12.95-15.50) | 0.596 |
| CRP (mg/dL) median (IQR) | 10.6 (5.6-20.26) | 8.03 (4.01-14.9) | 13.49 (8.91-28.98) | 0.000 |
| Procalcitonin (ng/mL), median (IQR) | 0.13(0.08-0.38) | 0.10 (0.06-0.28) | 0.25 (0.12-0.47) | 0.000 |
| IK6(pg/mL), median (IQR) | 42.2 (22.1-111.65) | 31.05 (9.90-42.35) | 56.55 (21.88-188) | 0.001 |
| Ferritin (ng/mL), median (IQR) | 1118(567-2052) | 927(398-1398) | 1416 (805-2606) | 0.007 |
| Troponin (pg/mL) , median (IQR) | 12.85(10.69-18.05) | 15.53 (10.25-17.23) | 12.85 (9.29-123.08) | 0.945 |
| SaO2/FiO2, median (IQR) | 424(346-448) | 433(419-452) | 361(180-428) | 0.000 |

PT: prothrombin time; aPTT: activated partial thromboplastin time; IQR: Interquartile range; LDH: Lactate Dehydrogenase; CRP: C-reactive protein; IK6: Interleukin 6.
Normal range: PT (11 – 13.5 seconds); aPTT (27- 40 seconds); Fibrinogen (130 – 400 mg/dL); D-dimer ( $< 0.5 \mu\text{g/mL}$ ); Platelet count ( $150 \times 10^9/\text{L} - 400 \times 10^9/\text{L}$ ); Lymphocytes count ( $1.2\text{-}3.5 \times 10^9/\text{L}$ ); Neutrophils count ( $1.4\text{-}6.5 \times 10^9/\text{L}$ ); LDH (135-225 U/L). CK (34-145 U/L); CRP ( $< 0.5 \text{ mg/mL}$ ); Creatinine: (0.5-0.9 mg/mL); Bilirubin (0.15-1.2 mg/mL); Albumin (3.5-5.2 gr/dL); Procalcitonin (0-0.5 ng/mL). IK6 (0-3.4 pg/mL). Ferritin (15-150 ng/mL). Troponin (0-210 pg/mL).

The table 3 includes the areas under the curve (AUC) for the features with significant differences between both nCG and CG groups as well as the cut-off points for the parameters that maintained their statistical significance in the logistic regression analysis. These variables were the SOFA score ≥2 (70% sensitivity and 76 % specificity) and CRP ≥9.1 (75% sensitivity and 53% specificity).

**Table 3.**
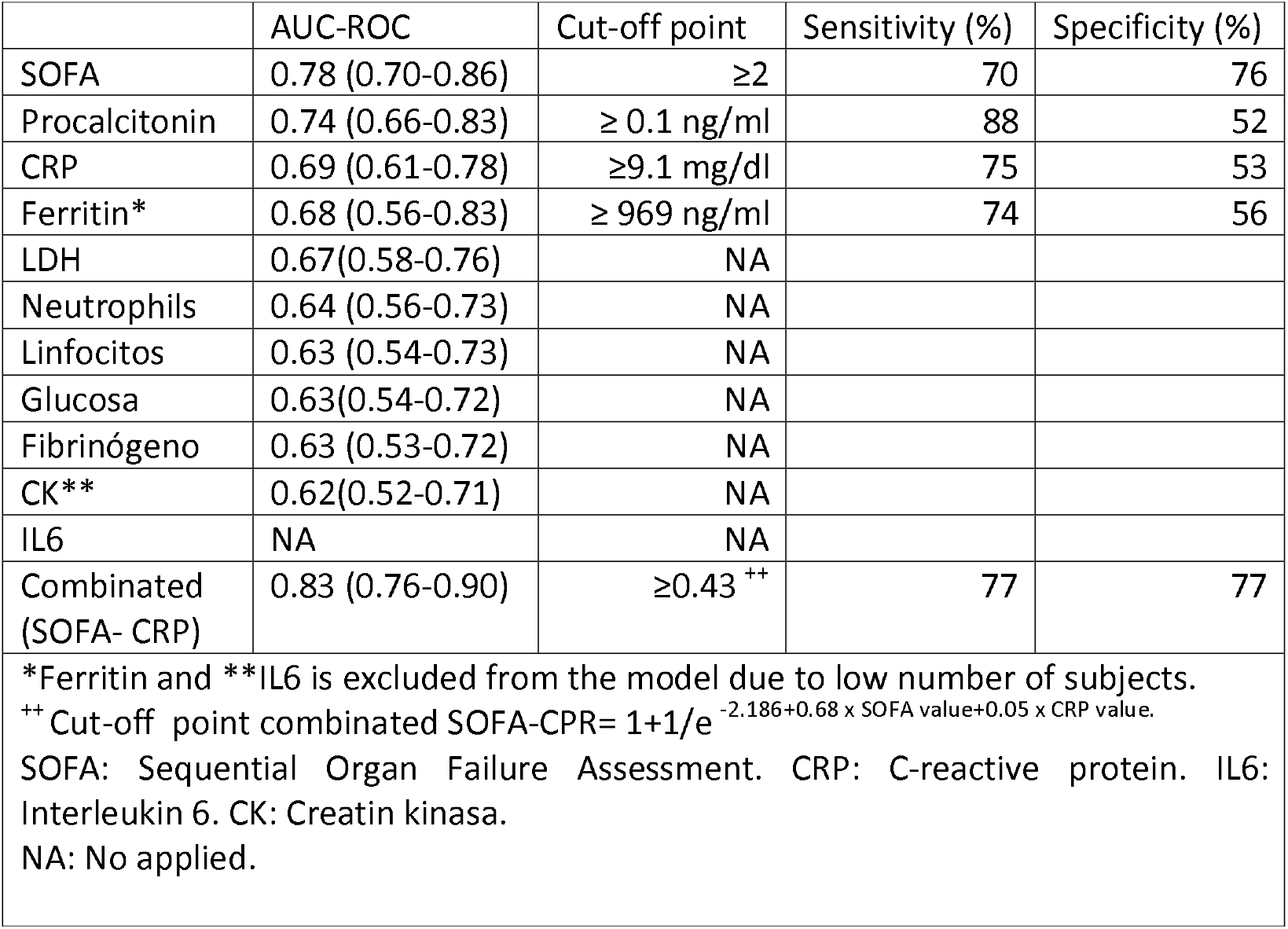
AUC-ROC and cut-off point, sensitivity and specificity of parameters of COVID-19 patients at the time of admission for the probability of critical care.

A multivariable logistic regression model was performed and the high SOFA score resulted in an odd of requiring admission at the CG of 1.968 as well as a high CRP level at admission associated with an odd of critical care need of 1.052 (Table 4). The presence of high levels of procalcitonin resulted also in an odd of 1.152 for the need of critical care, but it was finally removed because of the low specificity reported in the area under the curve. Moreover, the figure 1 represents the ROC curves for the SOFA score as isolated variable and SOFA score plus CRP levels showing a significant difference when both SOFA and CRP were linked reaching a sensitivity and specificity levels of 77%. The ROC area here observed (0.8) suggests the high predictive value of this model when considering the SOFA score and CRP at the moment of admission at the hospital.

**Figure 1.**
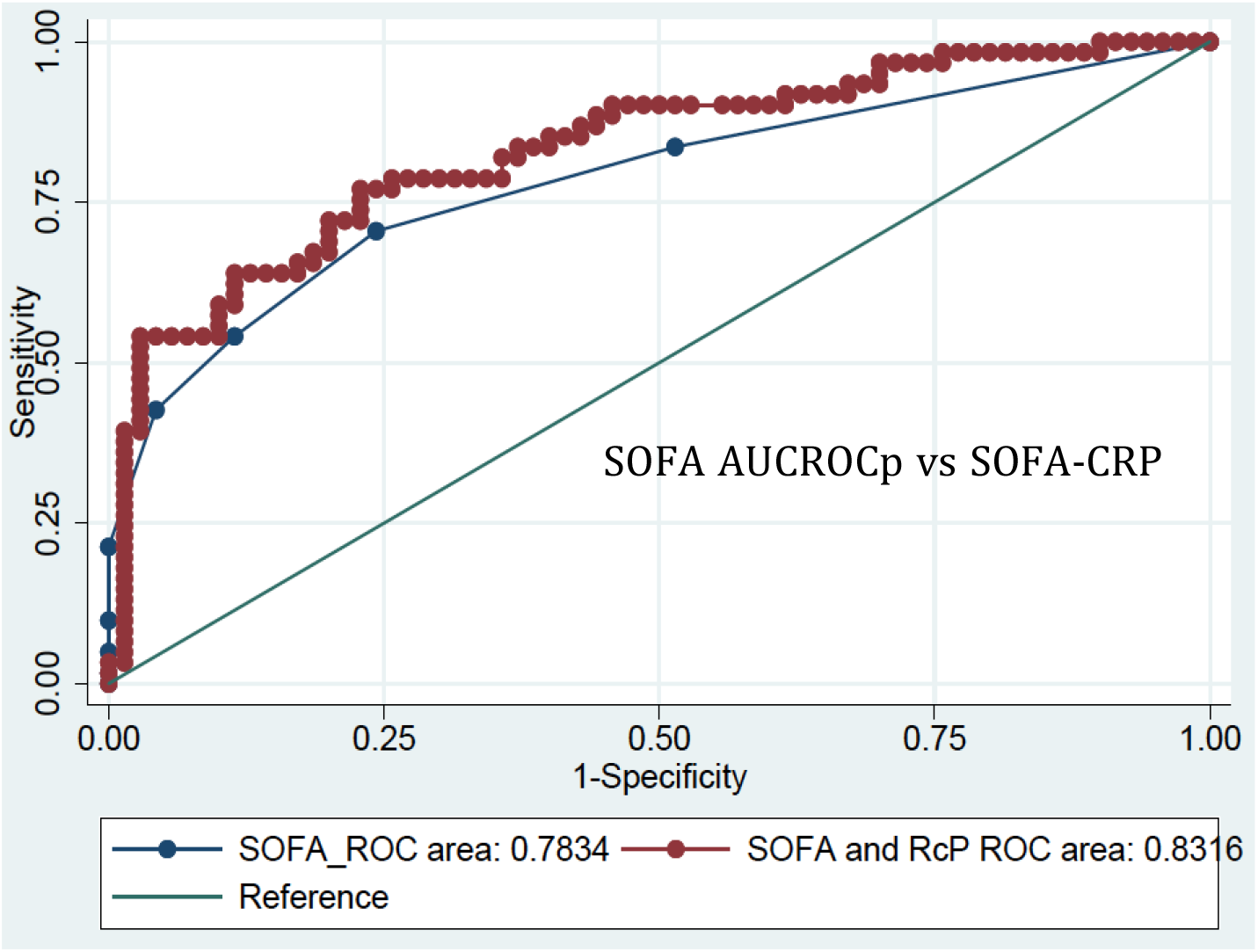
AUC-ROC difference between SOFA and combinated SOFA-RCP.

**Table 4.**
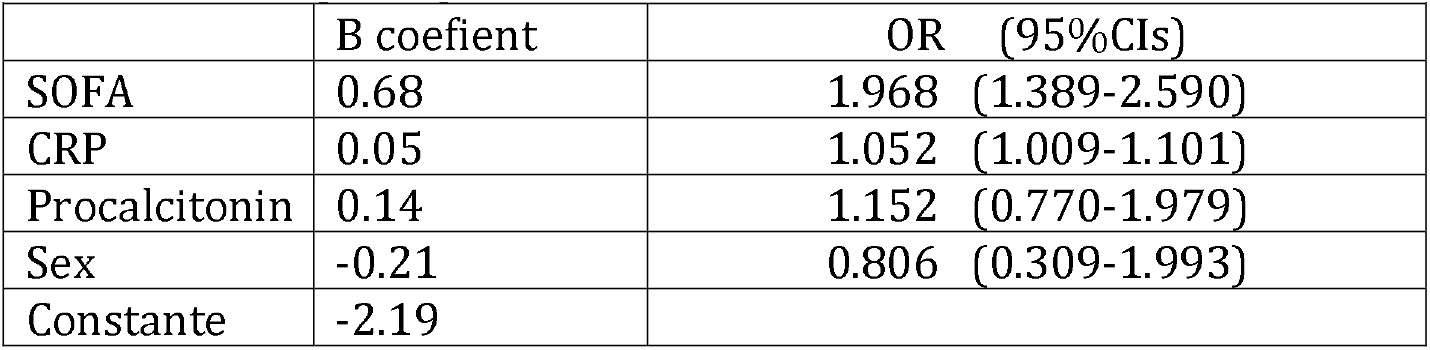
Multiple regression with the 4 covariates of interest.

Table 5 represents the distribution of patients in both non-CG and CG groups according to the inflammatory profiles upon arrival at the hospital and of note, 67.7% of patients with an inflammatory profile 1 (CRP> 9.1 mg / dl and ferritin> 969 ng / ml) at admission required critical care whilst only 16.1% presenting at admission with an inflammatory profile 4 (CRP <9.1 mg / dl and ferritin <969 ng / ml) required critical care.

**Table 5.**
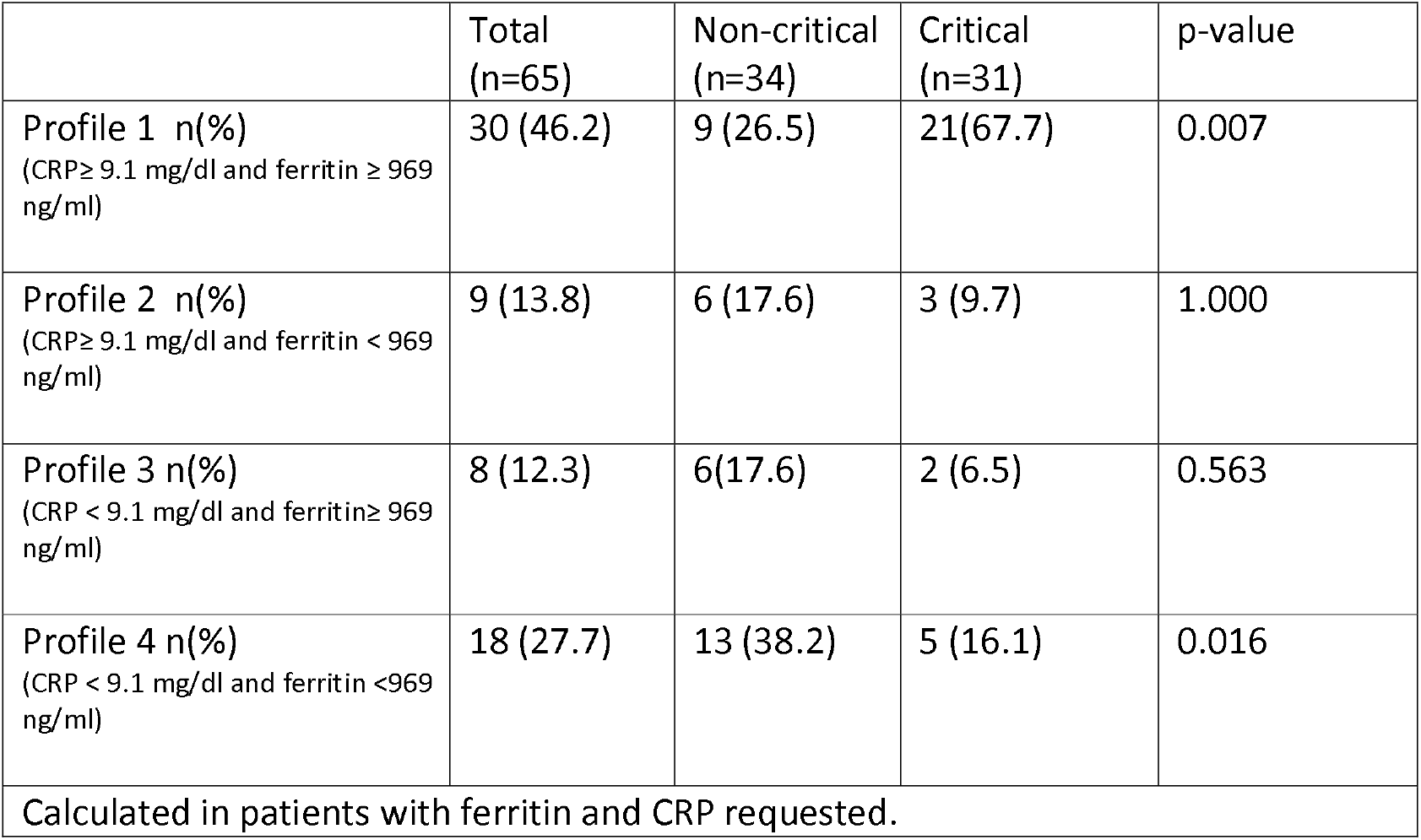
Profile parameters of inflammation of COVID-19 patients on admission according to the destination upon admission.

|  | Total<br>(n=65) | Non-critical<br>(n=34) | Critical<br>(n=31) | p-value |
| --- | --- | --- | --- | --- |
| **Profile 1 n(%)**<br>(CRP≥ 9.1 mg/dl and ferritin ≥ 969 ng/ml) | 30 (46.2) | 9 (26.5) | 21(67.7) | 0.007 |
| **Profile 2 n(%)**<br>(CRP≥ 9.1 mg/dl and ferritin < 969 ng/ml) | 9 (13.8) | 6 (17.6) | 3 (9.7) | 1.000 |
| **Profile 3 n(%)**<br>(CRP < 9.1 mg/dl and ferritin≥ 969 ng/ml) | 8 (12.3) | 6(17.6) | 2 (6.5) | 0.563 |
| **Profile 4 n(%)**<br>(CRP < 9.1 mg/dl and ferritin <969 ng/ml) | 18 (27.7) | 13 (38.2) | 5 (16.1) | 0.016 |
| Calculated in patients with ferritin and CRP requested. |  |  |  |  |

## Discussion

In this retrospective study, we report that the presence of C-reactive protein levels ≥ 9.1 mg/dl and SOFA score ≥2 in patients COVID-19+ at the moment of admission at the hospital are independent predictors for requiring critical care unit admission with a sensitivity and specificity levels of 77%. To date, there are not literature regarding both markers together in order to predict intensive care admission in COVID-19 patients.

The SOFA score and its association with severity in patients COVID-19 has been previously reported in other studies (5,6). However, the score of 2 reported in our series resulted into an AUC of 0.78 (0.76-0.86) what is acceptable and, in fact, it is the cut-off considered for distinguishing between septic and non-septic patients (16). Although this is applicable to bacterial infections, viral infections can also result in sepsis syndrome and nearly 40% of adults with community-acquired pneumonia due to viral infection develop a sepsis syndrome (22).

Furthermore, in our series the role of SOFA score as predictor of the need for critical care was sustained after correction for the other main covariates like CRP, procalcitonin and sex.

As far as CRP is concerned, high levels and its association with prognosis and severity in COVID 19 disease has been reported (23, 24). In our series, the role of CRP as predictor of critical care need maintains its significance in the univariate analysis and the cut-off of 9 mg/dl showed to be the most sensitive and specific factor to discriminate patients who will require critical care versus those who will not. Its role as predictor was maintained in the multivariate analysis. As its synthesis mainly occurs in response to pro-inflammatory cytokines (10), most notably IL-6 and to a lesser degree IL-1 and tumor necrosis alpha (TNF-α), its determination is recommended at the moment of admission and is more accessible than the other cytokines levels determination.

Combining both CRP and SOFA, we found in our series an AUC of 0.83 (0.76-0.90), what is excellent for predicting intensive care at admission and, in our knowledge, had not been previously reported. However, its biological plausibility is evident because, as it happens in other infectious processes (25), the combination of one of the main inflammation markers (CRP) with a validated organ failure scale in the infectious context (SOFA) is able to provide more accuracy in the diagnosis/prognosis of this disease.

Another relevant finding in this study has been than almost 70% of patients presenting at admission with CRP higher than 9.1 mg/dl and ferritin higher than 969 ng/ml required critical care suggesting their accuracy for predicting prognosis. On the other hand, the inflammatory profile with CRP less than 9.1 mg/dL and ferritin less than 969 ng/ml would help to identify patients with low probability of requiring critical care and, in fact, only 16% of patients in our series with this profile required admission into the CG.

As it has been previously mentioned, the CRP is an intermediary of the IL-6 track. Interleukin (IL) −1 and IL-6 have all been shown to trigger acute endothelial cell activation (24) resulting in high levels for these and another cytokines in critically ill patients (26). Ferritin is also an intermediary of the IL-1 track (10,11) so the use of both CRP and ferritin will give us an inflammation pattern that can be important with prognostic impact.

Procalcitonin has been related with prognosis in patients with inflammatory response similar to what we found in our series (27). However, its role as predictor for CG was not maintained in the multivariate analysis maybe because of its poor accuracy for the diagnosis of viral infections (28).

Sex has been described in other series as a prognostic factor (8, 9) but for overall mortality and in our study, being male did not predict higher risk for requiring critical care. The same effect occurred for the time until either admission in the hospital or start treatment. Underlying disease comorbility did not differentiate upon arrival at the hospital between both groups with exception of asthma which has been previously identified according to other works (29).Concerning treatments, more patients in the critical group received high-dose heparin, interleukine-6 receptor inhibitors and corticosteroids what it is expected because of their severity.

LDH, CK and White series appear to be prognostic factors in other publications (3,5,7,30), and specifically related to viral load (LDH and CK) (30). In our case, there were not included in the multivariate analysis for presenting AUC ROC lower than suitable for diagnosis.

Our study has some limitations, because of the retrospective nature as well as because some laboratory tests were missing in some patients.. Although the sample size is rather small, the inclusion of randomly adult patients COVID-19 positive is representative of the number of cases treated in critical care units.

Finally, overall and as expected, patients that required admission into the critical care units had a significantly higher inpatient mortality rate, more than twice, compared to those who did not require it. The mortality rate here reported is consistent with that reported in other series (3) and based on this fact, it results crucial the potential identification of this group of patients at the moment of admission. As patients presenting with a SOFA score >2 plus high levels of CRP will more likely require critical care, further actions could be implemented in order to reduce the requirement of critical care in these patients with the final objective of decreasing the inpatient mortality rate.

## Data Availability

The authors confirm that the data supporting the findings are available within the article and its supplementary material

## Acknowledgments

Weacknowledge to Maria-Victoria Mateos from Hematology Department for her contribution with the review of this manuscript. Our appreciation to Pedro Luis Sánchez-Fernández from Cardiology department as well as to all doctors and healthcare personn involved in the COVID-19 Working Team.

We also thank to all members of Department of Anesthesiology and Réanimation: Jordana Almeida, Daniel Álvarez, Laura Alonso, Begoña Alonso, Marta Criado, Isabel de Celis, Alberto de Diego, Esther del Barrio, Carlos Espinel, lolanda Freire, Emilio Garcia, Isabel Garrido, Eugenia González, José Luis González, María Heredia, Felipe Hernández, Janna Herrmmanova, José Luis Iglesias, Elisa Jausoro, Diego Leoz, Adela López, Rocío López, Pamela López, Virgilio Martín, Laura Nieto, M Jesús Pascual, Diego Pérez, Isabel Pingarrón, Rosa Prieto, JM Rodríguez, Mercedes Rojo, Cruz Ruano, Daniel Salgado, Lucio San Norberto, Pilar Sánchez, Manuel J Sánchez, Eduardo Sánchez, Francisco J Sánchez, David Sánchez, Valentín Santana, Ignacio Trejo, Joaquín Valdunciel, Ana Vara, M José Villória, Gema Yusta.

We also thank to all COVID-19 patients admitted at University Hospital of Salamanca as their families.

